# The SCAMP Research Challenge programme: Co-developing research with young people for young people

**DOI:** 10.64898/2026.09.15.26363137

**Authors:** Nicole Curtis, Rhiannon Thompson, Lan Cheng, Chen Shen, Rachel B Smith, Rakhi Biswas Evans, Martina Di Simplicio, Mireille B Toledano

**Affiliations:** Mohn Centre for Children’s Health and Wellbeing, School of Public Health, Imperial College London, UK; Department of Epidemiology and Biostatistics, School of Public Health, Imperial College London, UK; MRC Centre for Environment and Health, Department of Epidemiology and Biostatistics, School of Public Health, Imperial College London, UK; National Institute for Health Research School of Public Health Research, Imperial College London, UK; National Institute for Health Research Health Protection Research Unit in Radiation Threats and Hazards, Imperial College London, UK; National Institute for Health Research Health Protection Research Unit in Chemical and Radiation Threats and Hazards, Imperial College London, UK; Division of Psychiatry, Department of Brain Sciences, Imperial College London, UK

## Abstract

**Background:** Public and participant involvement (PPI) is central to conducting ethical, relevant and impactful research. In adolescent mental health research, involving young people is particularly vital given their unique lived experiences, perspectives and unmet mental health needs. The SCAMP Research Challenge (RC) was developed to embed meaningful and reciprocal youth participation into the Study of Cognition, Adolescents and Mobile Phones (SCAMP), the world’s largest cohort study on adolescent mobile phone use, cognition and mental health.

**Objectives:** The RC sought to expand cohort recruitment through peer-led approaches, engaging students as active partners in co-delivering data collections within their schools, whilst providing training to support the development of their own research proposals using SCAMP data. Selected projects were subsequently taken forward for further collaborative work with students.

**Methods:** Across two academic years (2022–2024), the year-long RC programme was delivered twice to 19 secondary schools in Greater London. Each school nominated a student RC team, which collaborated with SCAMP researchers to lead peer recruitment campaigns, co-organise school-based data collection sessions and develop their own research proposals. Selected teams advanced their projects through structured workshops in literature review, data interpretation and research communication. Engagement and recruitment outcomes were compared with previous SCAMP recruitment and data collection waves.

**Results:** RC teams successfully recruited 1,982 new participants into the SCAMP cohort, achieving higher participation rates within shorter timeframes than prior researcher-led approaches. Students demonstrated strong engagement by leading recruitment, facilitating data collections and co-developing seven research projects exploring topics such as digital technology and social media use, digital wellbeing and mental health. All projects are currently being prepared for submission to peer-reviewed journals. Participants reported increased confidence, communication and analytical skills, with several continuing to engage as SCAMP Research Ambassadors or members of the Young People’s Advisory Group (YPAG).

**Discussion:** The RC illustrates the feasibility and value of integrating PPI and participatory methods into large-scale youth research. Peer-led recruitment proved highly effective for enhancing engagement and data quantity, while the co-development of research projects fostered skills, empowerment and sustained involvement. Challenges included potential opportunity bias due to teacher nominations and inconsistencies in school recruitment logistics. Future iterations should employ transparent, standardised recruitment to ensure inclusivity and equity.

**Conclusion:** The SCAMP Research Challenge demonstrates that co-developing research *for* and *with* young people can strengthen scientific outcomes while empowering youth as co-contributors. Embedding participatory models in educational settings offers a scalable approach to improving research relevance, impact and youth engagement in public health research.

## Introduction

Recent years have seen a marked increase in patient and public involvement (PPI) in child and adolescent mental health research, with approaches ranging from advisory roles to more collaborative models of co-production.[1] Evidence suggests that involving young people can enhance relevance, acceptability and ethical integrity of research, particularly when they contribute to shaping research questions, design and development.[2] However, the literature highlights considerable heterogeneity in how PPI is implemented and evaluated, with advisory groups being the most common approach albeit often limited in depth and meaningful decision-making processes.[3] Further, while emerging research has begun to highlight the effectiveness of co-production, there is still limited evidence demonstrating the impact of PPI on research outcomes.[4] Qualitative research indicates that effective adolescent involvement should extend beyond procedural inclusion to encompass trust, shared decision-making and empowerment through skill development. Overall while adolescent PPI is increasingly part of mental health research, key gaps remain in the efficacy of approaches, transparency of reporting and evaluation of impact.[2]Young people’s mental health problems are on the rise in the UK with existing systems struggling to meet the demand.[5, 6] While adolescence is a crucial time for mental health, with most lifetime disorders developing during this period of significant social, emotional and neurodevelopmental change, modern day adolescents face a unique set of challenges, including the impact of digital technologies and broader societal challenges which can shape mental health risk and resilience in complex ways.[7, 8] Much is still not understood about the risks and protective factors contributing to young people’s mental health. Further, adolescents also lack decision making power, as key decisions about their physical and mental health are ultimately governed by adults and in society, where they are widely disenfranchised. [8, 9] This considered, adolescent mental health research should not only be methodologically robust, but relevant, inclusive, responsive, and empowering, ensuing young people’s perspectives are central to the development of effective interventions and services.[8]

The Study of Cognition, Adolescents and Mobile Phones (SCAMP) is the largest study in the world examining adolescent mental health, cognition and mobile phone use, with over 12,000 students participating across Greater London.[10] Over the last 11 years, this cohort has provided a rich dataset, enabling researchers to answer questions about digital technology and social media use, sleep, COVID-19, multilingualism, air pollution, noise exposure, greenspace and physical activity, with a focus on mental health and cognition.[11–18] Importantly, SCAMP has a strong history of public involvement, including a Young People’s Advisory Group (YPAG), Research Ambassadors and initiatives for parents from seldom-heard communities. These engagement endeavours have supported study design, materials development, recruitment methods and data analysis reviews. However, consistent with the wider literature, much of this involvement has centred on consultive roles, reflecting broader challenges in moving towards sustained, impactful co-production and in evidencing the added value of youth involvement beyond specific study concepts.[1, 4]

Following the COVID-19 pandemic, initial recruitment attempts yielded low school response and participation rates, likely reflecting the ongoing disruption and hesitancy following the pandemic school closures.[20, 21] In response to both the field-wide gaps and practical recruitment challenges, SCAMP researchers developed the Research Challenge (RC), a novel reciprocal programme integrating public and community involvement, engagement and participation. Designed to recruit older adolescents (16–18-year-olds) through opt-in consent, the RC moved beyond traditional PPI structures, by embedding young people as active contributors to the research process through peer-led recruitment, in-school data collections and the co-development of research proposals using cohort data. The programme sought to strengthen research impact and give back to schools and students by enhancing young people’s scientific understanding, communication and research skills, as well as their overall confidence. The RC was also explicitly designed to align with the Gatsby Benchmarks, supporting schools to meet national careers curriculum requirements. Specifically, the main objectives were to (1) expand cohort recruitment through peer-led methods, (2) deliver in-school data collection sessions, (3) support students in developing research proposals using SCAMP data and (4) collaborate further with students on selected projects.

## Methods

### School and Participant Recruitment

Following a pilot study which assessed programme feasibility, a full-scale Research Challenge (RC) programme was rolled out in September 2022. Schools were eligible to participate in the programme if they met the following criteria: they were located within Greater London, taught Years 12 and 13 (students aged 16 to 18), could provide suitable facilities for data collection (adequate ICT provisions for online assessments and space for biosample data collection), were willing to nominate a student team and staff liaison to support with the delivery of the programme and complied with study timelines and requirements. For Year 2 of the programme (2023-24), schools were required to have at least 30 students willing to complete the online assessment; schools with fewer than 30 students across both year groups were included if all students participated. Students were eligible to participate if they met the following requirements: they were enrolled in Years 12 or 13 at participating schools, could provide informed consent and complete the online assessments, with reasonable adjustments if needed.

In Year 1 (2022–23), school recruitment was conducted via cold calling, emails and in-person visits. Once schools and the students agreed to participate, teachers were asked to nominate up to eight Year 12 students to form a RC team. Once selected, both the students and teachers were given further information on the programme’s structure, roles and expectations. This included the RC team’s responsibility to help co-organise and recruit their peers to participate in a data collection at their school as well as develop their own research question. In Year 2 (2023–24), recruitment followed the same approach but also drew on collaborations with the North East London NHS Foundation Trust, an NHS Clinical Research Network, which helped facilitate access to additional schools and staff to support during school visits. While most schools in Year 1 engaged fully with both data collection and research, Year 2 introduced flexibility, with some schools participating only in the data collection, while others participated in the full programme.

### Kick offs

Once RC teams were established, SCAMP researchers delivered kick-off presentations at each school. These included an introduction to the SCAMP study, an outline of the RC programme, and training in engagement, organising a school-based data collection, and how to develop research questions. Students were taught key epidemiological concepts and methods to help them understand how these related to the data collection, as well as what to consider when designing their recruitment campaigns within their schools. After the sessions, RC teams were provided with handbooks, presentation slides from the kick off presentations, data collection visit forms and research question submission forms, as well as group and individual feedback forms. Teachers were also provided with individual feedback forms.

In Year 1, kick-off sessions were delivered in person and deadlines for returning forms were fixed (e.g., data collection forms due two weeks before visits; research question forms by March). In Year 2, following the feedback from the previous year, in-person kick-offs were supplemented with virtual check-ins to provide ongoing support.

### School-based data collection

Data collection was co-organised by RC student teams, teachers and SCAMP researchers. The RC teams completed data collection visit forms specifying proposed dates and times, available space within school (e.g., sports hall, computer room, classrooms), numbers of students and the chosen Tier of participation (see Table 1). The Tier choice required teacher input on behalf of the school, to ensure suitability and available resources.

**Table 1.**
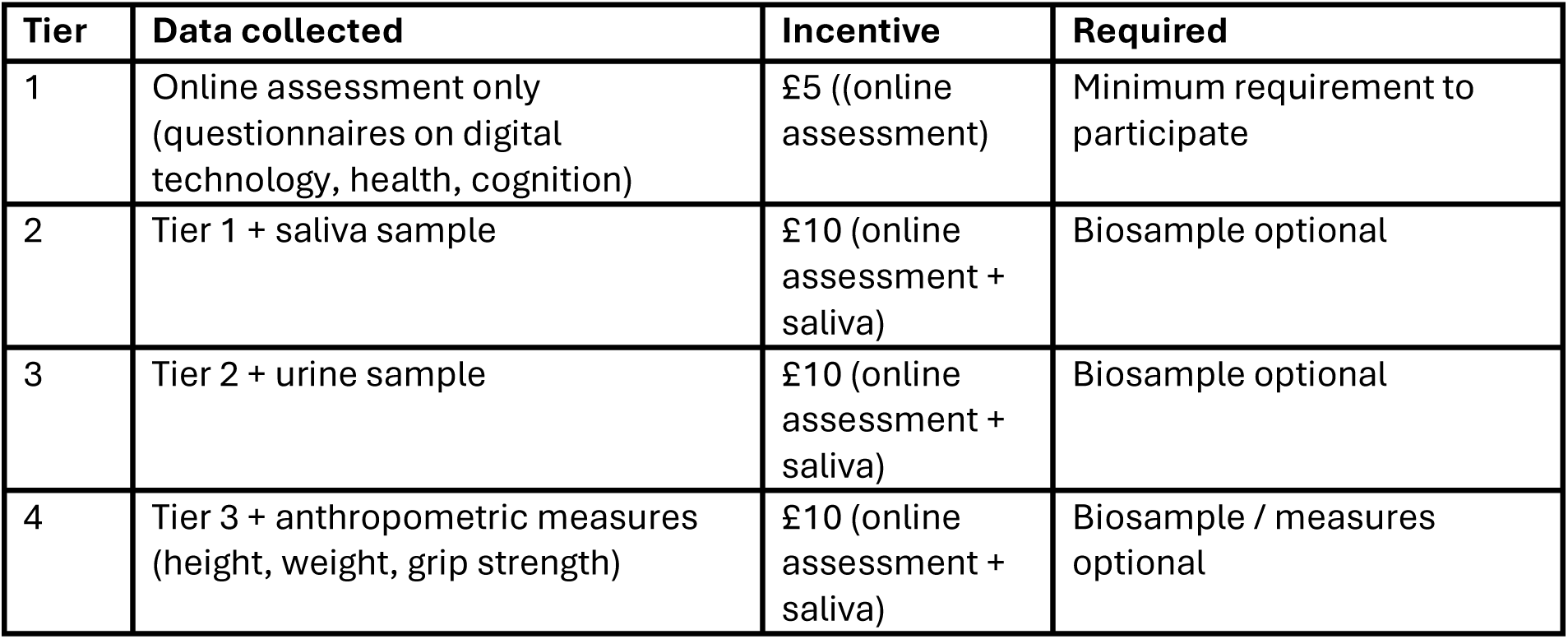
Data collection tiers available to participating schools.

| **Tier** | **Data collected** | **Incentive** | **Required** |
| --- | --- | --- | --- |
| 1 | Online assessment only<br>(questionnaires on digital technology, health, cognition) | £5 ((online assessment) | Minimum requirement to participate |
| 2 | Tier 1 + saliva sample | £10 (online assessment + saliva) | Biosample optional |
| 3 | Tier 2 + urine sample | £10 (online assessment + saliva) | Biosample optional |
| 4 | Tier 3 + anthropometric measures<br>(height, weight, grip strength) | £10 (online assessment + saliva) | Biosample / measures optional |

Participants received vouchers as incentives for completing the online assessment (£5), as it captures essential data on health, mental health, digital technology use and cognition, and for providing the saliva sample (£5), which enables genotyping analysis. These elements are the core components of the SCAMP study. Data collection was led and supervised by SCAMP researchers under exam-like conditions, with biosamples collected by trained staff and selected anthropometric measures collected by trained RC students under supervision.

In RC Year 1, in-school data collections occurred between November 2022 and February 2023. In RC Year 2, data collection was extended from November 2023 to June 2024 to accommodate both full- and partial-participation schools. In both years, RC teams played key roles in planning, recruiting peers (often via assemblies and promotional materials) and supporting logistics on the day.

### Research question selection and development

After completing the data collections, RC teams submitted their research question forms, which guided them through formulating a hypothesis, defining exposures and outcomes and considering confounders. Students were informed during the kick-off sessions that, while all proposals would receive feedback, only a select number of projects demonstrating feasibility and alignment with SCAMP data would be taken forward for further analysis and dissemination. Researchers assessed submissions against six criteria (novelty, importance, reasoning, variable choice, confounder consideration, effort). Based on the highest-rated submissions, three projects were selected in Year 1 and four were chosen in Year 2. Across both years, selection was announced in March, after which winning teams progressed to the research stage.

### Awards ceremony

Each year an Awards Ceremony was hosted at Imperial College London, to recognise and celebrate the RC students’ achievements. RC student teams, teachers and SCAMP researchers attended, as well as key stakeholders such as scientists and clinicians from the Mohn Centre for Children’s Health and Wellbeing, Imperial’s School of Public Health, University College London and the National Institute for Health and Care Research. The RC teams were awarded certificates for participation, whilst select teams received awards for excellence in data collection (e.g., ‘highest overall number of participants’, highest online assessment completion rate’, ‘most saliva samples’) and recognition of outstanding research proposals. In Year 1, the ceremony was held in March 2023, marking the end of the data collections and the transition into the research phase. In Year 2, the ceremony took place in July 2024, after the final data collection visits, the ceremony included presentations by Year 1 winners on their research projects. Both events included guest lectures from scientists and industry partners, as well as an ‘Expert Panel QCA’, during which the RC teams could engage with experts from the public health sector to learn more about their careers and research. The events were held after school hours to ensure maximum attendance and allow for commuting time.

### Research projects

The winning RC teams worked with researchers from March onwards to develop their projects through structured training, including how to conduct literature searches, write reviews and plan analyses. Teams received feedback on literature searches and reviews, and prepared data analysis plans which were refined during in-person workshops using dummy datasets in R. Due to data restrictions, final analyses were conducted by researchers but discussed collaboratively with the teams. Researchers were available to support the RC teams when needed with frequent check-ins and provided feedback at each stage of the project development. Data analysis training was held in August in both Year 1 and 2, and projects were further developed with researcher support into the following academic year. See Figure 1 for a timeline of the RC programme across both years.

**Figure 1.**
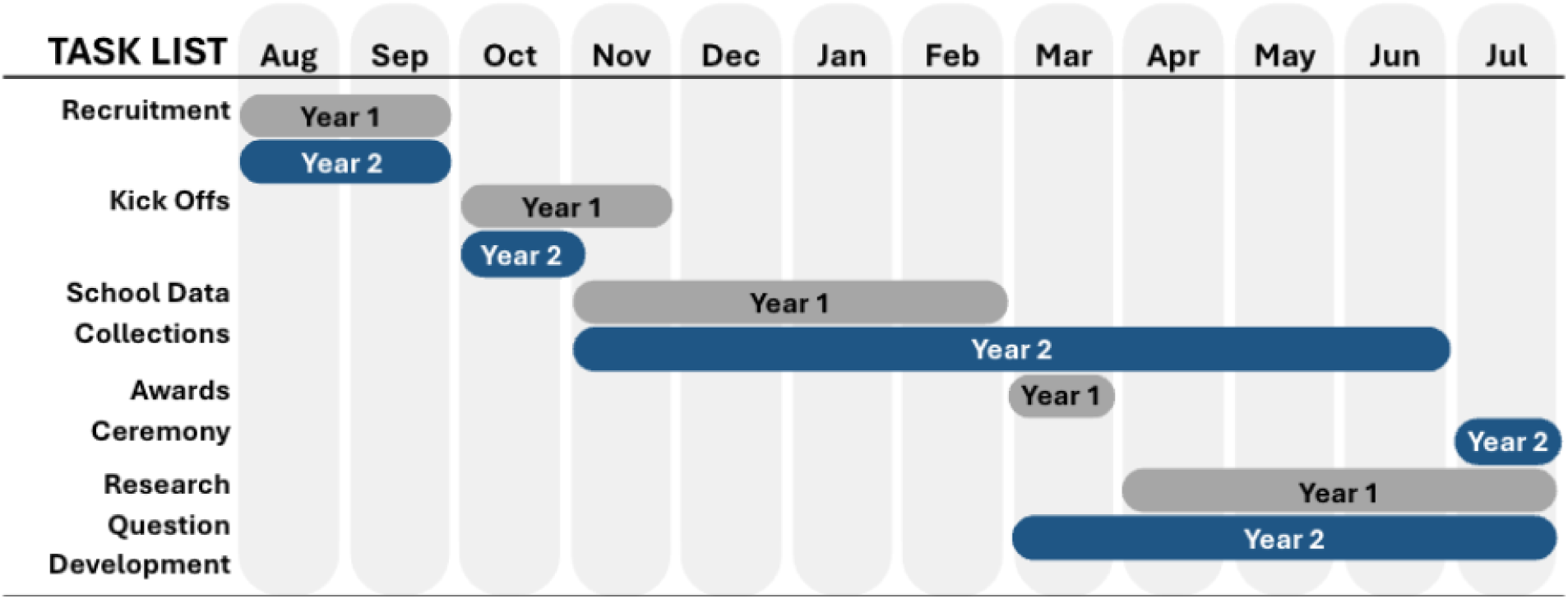
Timeline of SCAMP Research Challenge Years 1 and 2.

### Ongoing engagement

Following both years of the RC, researchers continued to support student teams in refining their projects and reviewing final analyses. Students were invited to remain engaged through structured opportunities, including joining SCAMP’s YPAG, the Research Ambassador programme, undertaking work experience with SCAMP or contributing to public engagement activities. For Year 3 (2024-25) of the RC, a new cohort of 20 RC students from participating SCAMP schools were onboarded to, using the findings from the RC Year 1 and 2 projects and their lived experiences, co-design a peer-led intervention addressing digital technology use and adolescent wellbeing in schools. This collaboration resulted in the development and delivery of the ‘Scroll Smart Study (SSS)’, which commenced in Winter 2025–2026.

### Ethics

The North-West Haydock Research Ethics Committee approved the SCAMP study protocol and subsequent amendments (ref 14/NW/0347). All participants in the data collection were provided with information sheets, completed consent forms and were informed that they could withdraw from the study at any time. The study was conducted in accordance with the Declaration of Helsinki.

## Findings

### School recruitment

SCAMP researchers contacted a total of 186 schools across both years of the RC (109 in Year 1 and 77 in Year 2). In Year 1, 12 schools agreed to participate, and 12 RC student teams were subsequently established, with each school participating in the entire programme and student teams leading peer recruitment. In Year 2, participation increased to 15 schools. Due to the revised programme structure, 11 student teams were formed whilst the four remaining schools opted to only take part in the data collection phase. Figure 2 shows a breakdown of school and student participation rates. All participating schools were small-sized secondary schools (<700 students), as shown in Table 2.

**Figure 2.**
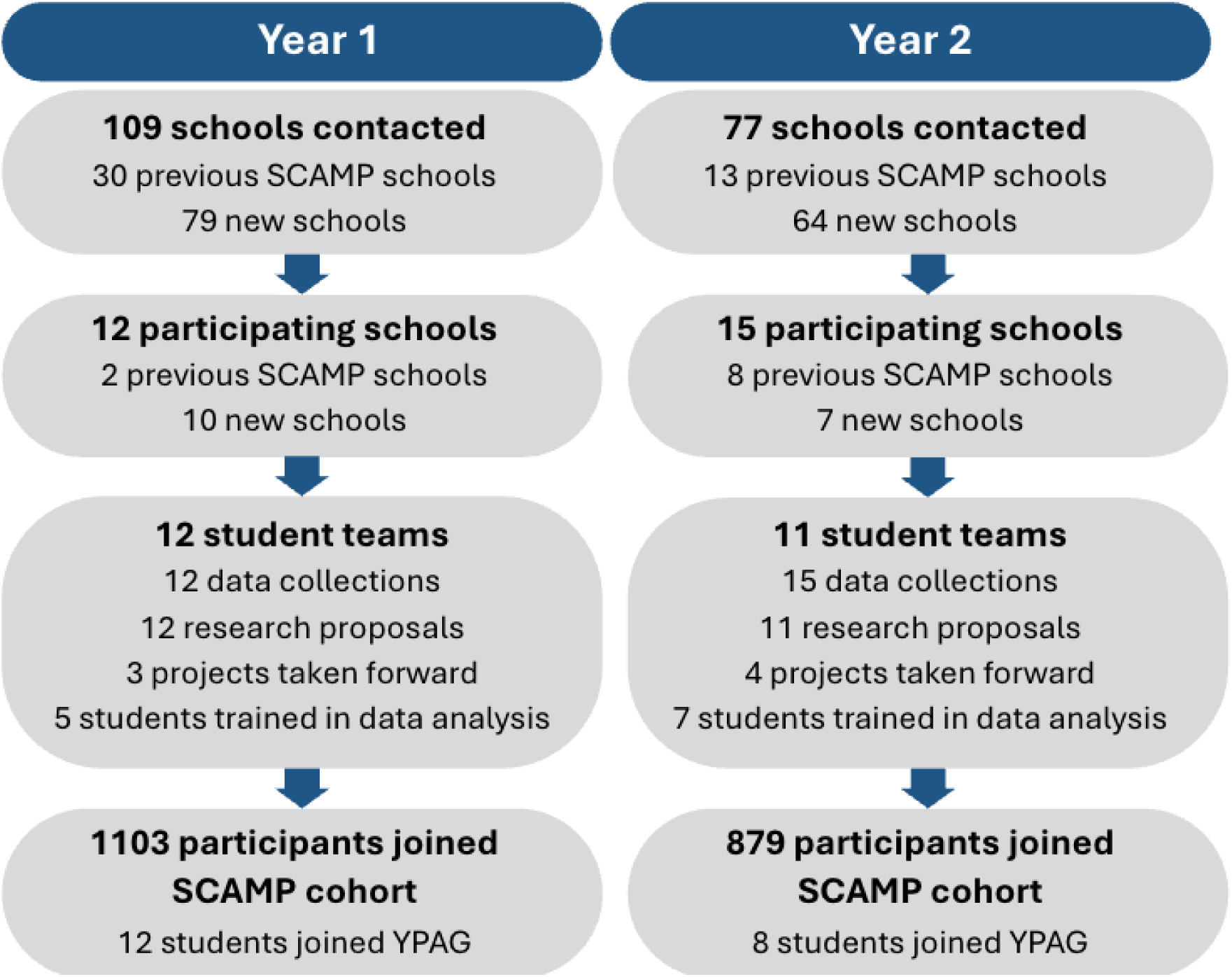
School and student recruitment and participation.

**Table 2.** School and student profile across all participating schools.

| **School** | **Location**<br>**(Borough,**<br>**Region)** | **Participation**<br>**Year** | **Sixth Form**<br>**Gender**<br>**Distribution** | **% Eligible**<br>**Free School**<br>**Meals (FSM)** |
| --- | --- | --- | --- | --- |
| **Ada Lovelace**<br>**CofE School** | Westminster,<br>Central London | Year 2 | Mixed | 23% |
| **Alec Reed**<br>**Academy** | Ealing (Northolt),<br>NWL London | Year 1 & 2 | Mixed | 41.3% |
| **Beal High**<br>**School** | Redbridge, NEL | Year 1 & 2 | Mixed | 20% |
| **Bishopshalt**<br>**School** | Hillingdon, NWL | Year 1 & 2 | Mixed | 26.6% |
| **Chislehurst**<br>**and Sidcup**<br>**Grammar**<br>**School** | Bexley, SEL | Year 1 & 2 | Mixed | ~3% |
| **City of London**<br>**Academy**<br>**(Southwark)** | Southwark, SEL | Year 1 | Mixed | 47.4% |
| **Claremont**<br>**High School**<br>**Academy** | Brent, NWL | Year 1 & 2 | Mixed | 18.2% |
| **Hammersmith**<br>**Academy** | Hammersmith &<br>Fulham, NWL | Year 1 | Mixed | 34.6% |
| **Islington**<br>**Collegiate**<br>**Sixth Form** | Islington, NEL | Year 1 & 2 | Mixed | Data not<br>public |
| **Marylebone**<br>**Boys' School** | Westminster,<br>Central London | Year 2 | Boys; mixed<br>sixth form | 49.3% |
| **Nower Hill**<br>**High School** | Harrow, NWL | Year 2 | Mixed | 18.9% |
| **Oaks Park**<br>**High School** | Redbridge, NEL | Year 1 | Mixed | 22.5% |
| **Pimlico**<br>**Academy** | Westminster,<br>Central London | Year 2 | Mixed | 57.5% |
| **St Angela's**<br>**Ursuline**<br>**School** | Newham, NEL | Year 1 | Girls | 39.7% |
| **Sydney**<br>**Russell School** | Barking &<br>Dagenham, NEL | Year 2 | Mixed | 27.3% |
| **The City**<br>**Academy,**<br>**Hackney** | Hackney, NEL | Year 1 | Mixed | 55.9% |
| **The Lady Eleanor Holles School** | Hounslow, NWL | Year 1 & 2 | Girls | Independent school – N/A |
| **Tiffin School** | Kingston upon Thames, SWL | Year 1 & 2 | Boys; mixed sixth form | 3.1% |
| **William Perkin CofE High School** | Ealing, NWL | Year 2 | Mixed | 25.1% |

The adjustments made in Year 2 were implemented in response to several factors: a reduction in class sizes compared to Year 1, which necessitated the inclusion of a greater number of schools to maintain a comparable participant sample size across both years; feedback from teachers and students indicating that offering the option to participate in only part of the programme could enhance school and student engagement; and considerations regarding the quality of research proposals and sustained involvement during the research phase, as allowing students to self-select into the full programme encouraged those with a genuine interest in developing research design experience to take part. This adaptive and collaborative approach proved instrumental in achieving one of the study’s main objectives, to recruit new participants into the SCAMP cohort, by allowing schools to engage at varying levels of involvement, thereby maximising participation across diverse educational settings. In these four data-collection-only schools teaching staff acted as the primary liaison between SCAMP researchers and the students.

The school recruitment strategies and outcomes varied across the two years, reflecting iterative refinements to the programme design. Overall, this adaptive approach appeared to enhance recruitment efficiency and geographical reach within Greater London.

### Kick offs

The kick-off sessions were delivered in person at each participating school across both academic years. After Year 1, feedback from the Year 1 RC students suggested that while the initial presentations were informative, they could benefit from a more interactive format. In response, the Year 2 kick-offs were redesigned to be more dynamic and accessible, incorporating interactive elements to enhance engagement and comprehension.

Further feedback from the Year 1 students indicated that greater support in between the kick-off and data collection sessions would be beneficial. Although the Year 1 students were informed that support was always available upon request, in Year 2, researchers introduced scheduled online check-ins with the student teams, which improved both the overall effectiveness of communication and time management. These refinements were positively received, with Year 2 students reporting a clearer understanding of their roles and improved coordination with the research team, as well as the overall research process. The enhanced communication structure appeared to strengthen school participation and sustain motivation, supporting the programme’s overarching aim of fostering meaningful student involvement in research and peer-led recruitment.

### School-based data collection

In Year 1, the data collection sessions took place between November 2022 and February 2023, with the RC student teams successfully recruiting 1,103 participants across the 12 schools. In Year 2, the data collection period was extended from November 2023 to June 2024 to accommodate both full- and partial-participation schools. During this period, an additional 879 students were recruited from the 15 participating schools. This resulted in the addition of 1,982 new participants to the SCAMP study, recruited from 19 different schools across Greater London.

The involvement of RC student teams was central to the success of the data collection sessions. Student teams led peer recruitment campaigns through assemblies, digital materials, and word-of-mouth, which contributed to high participation rates and efficient coordination during school visits. See Figure 3 for examples of student recruitment campaigns. This collaborative, student-led approach enhanced engagement among both students and staff and proved effective in expanding research participation through in-school data collection. Compared with previous SCAMP recruitment strategies, the peer-to-peer model achieved higher participation within a shorter timeframe, underscoring the value of student-led methods for improving recruitment efficiency. Overall, these findings demonstrate that actively involving young people in both peer recruitment and the practical delivery of research activities can significantly strengthen school-based research implementation.

**Figure 3.**
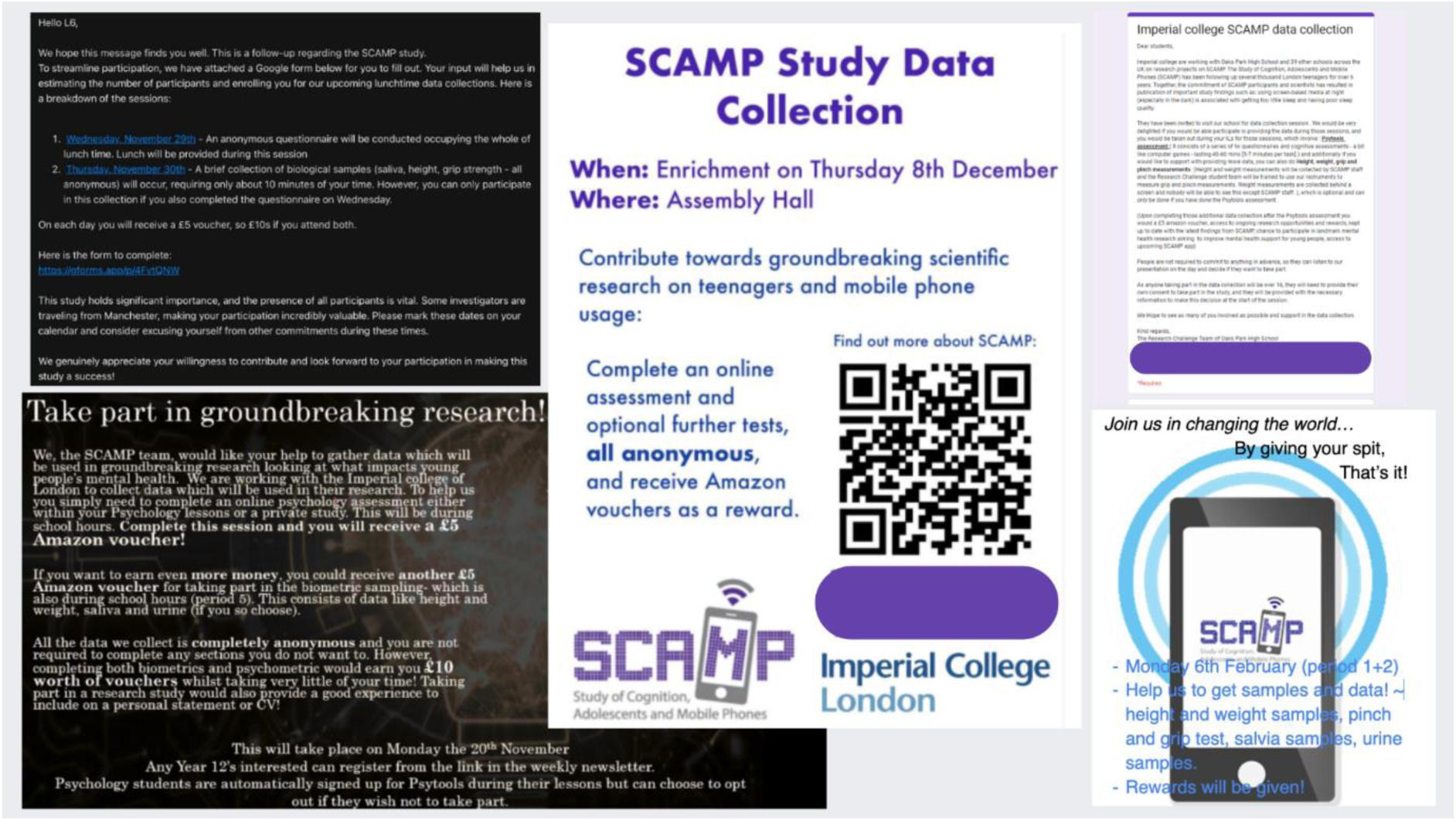
RC student recruitment campaigns.

### Research project development

Across both academic years, the RC student teams actively engaged with researchers to develop their research proposals, deepening their understanding of the research process from hypothesis formation to data interpretation. After a careful selection process, including feasibility within the constraints of the SCAMP dataset, a total of seven projects were chosen. In Year 1, the three selected projects explored (1) the relationship between videogaming and cognition, (2) the association between exposure to natural environments and academic performance as mediated by cognitive development, and (3) the effects of smartphone use duration on hand grip and pinch strength in adolescents. In Year 2, the four student proposals investigated (1) the relationship between scrolling on social media and emotion recognition ability, (2) associations between digital technology use and BMI, (3) the impact of academic performance on depression and anxiety symptoms, and (4) links between social media use and inattention in later adolescence.

Student participation during the research development stage varied according to interest and availability. Most students actively contributed to the question development, literature review and design stages, while fewer engaged in data analysis due to lower interest in quantitative methods and the training sessions taking place during the summer holidays. Attendance at the data analysis training increased from five of the RC students in Year 1 to seven in Year 2, and feedback on these sessions was consistently positive. Students highlighted the value of gaining practical research experience and developing transferable skills relevant to their final year of study and future educational plans. Both years concluded with well-attended and highly engaging award ceremonies at Imperial, which further celebrated students’ achievements and reinforced their connection with the research community.

### Ongoing engagement

Two projects from Year 1 and four from Year 2 are currently under development for journal submission, whilst the third project from Year 1 has now been published in a renowned academic journal. Additionally, 20 students across both years joined the SCAMP YPAG and have partaken in work experience placements at SCAMP, reflecting the sustained engagement and strengthening of ongoing partnerships between young people and SCAMP researchers. Further, the Scroll Smart Study, the pilot intervention co-designed with the 24-25 RC cohort, is nearing its end date. The peer-led intervention is being evaluated through a randomised control trial, with delivery of all four workshops now complete. One round of follow-up data collection is expected in May, after which data analysis, write-up and dissemination will commence.

### Student experience and feedback

A crucial part of the programme was ensuring RC student feedback was taken onboard and the recommended changes be implemented where possible; be it during Year 1, prior or during Year 2 roll out or noted for future iterations. The feedback collected via the feedback questionnaires was overwhelmingly positive, with some recommendations for future versions. RC students described the programme as “an amazing opportunity”, noting that they had “improved [their] communication skills and overall confidence”, whilst others indicated they had “gained skills in time management, collaboration, communication and organisation”. Several participants also reflected on how the experience influenced their future aspirations, with one student commenting that “the skills I learned will help me at university and in any career I pursue.”. Another participant and RC student team member continues to engage with SCAMP whilst currently reading medicine at Imperial. Further, many highlighted the importance of teamwork, describing how “our teamwork was strong, managing to delegate tasks and executing them effectively.”.

The constructive feedback received from Year 1 student teams focused predominantly on clearer guidance relating to research question development, more regular check-ins with researchers and increased hands-on student involvement during the data collection sessions. These aspects of the Challenge were addressed going into Year 2, with greater emphasis placed on making sure students were aware they could contact researchers at any time for support, alongside more frequent scheduled check-ins. Researchers also ensured student teams felt more involved during the data collection sessions, providing more training and assigning students from the RC teams roles prior to the visits. The suggested improvements offered by Year 2 student teams highlighted a desire for clearer team structures and increased opportunities for hands-on student involvement in research. This feedback will inform for future iterations of the programme. It has also been considered in the design of wider SCAMP research. For example, structured roles, regular researcher check-ins and greater opportunities for hands-on involvement was implemented throughout the development and delivery of the Scroll Smart Study.

Reflections for improvements considered, one of the most significant outcomes of the RC programme is the sustained involvement of select students beyond the original project cycle, with some continuing to be involved in the development of their projects to date as well as participation in the YPAG. This ongoing engagement reflects not only the motivation of these students to see their research through to completion and the capacity-building SCAMP offers but also underscores the impact and value of co-developing research *with* young people *for* young people.

## Discussion

This study explored a novel approach to co-developing mental health research *for* young people *with* young people. In line with the first objective, this peer-led approach resulted in the successful recruitment of nearly 2000 new participants to the SCAMP cohort across both academic years. This reflects the efficiency of leveraging students’ understanding of school culture and peer networks compared to previous researcher-led strategies. The integration of partnerships with the NHS Research Networks in Year 2 further enhanced access to eligible schools and supported a wider geographical reach, demonstrating the scalability of this participatory model. In line with the second objective, student teams effectively planned and facilitated data collection sessions in their schools. The third objective was achieved through structured guidance and collaboration, resulting in seven student-led projects covering topics ranging from social media use and emotion recognition to gaming, digital wellbeing and academic performance. Finally, the fourth objective was met through the advancement of selected projects in partnership with students, demonstrating the programme’s success in fostering engagement and providing practical experience in research design, data interpretation and collaborative effort. Several student-led projects have since progressed to further analysis and publication, as well as directly informed the student-led intervention, the Scroll Smart Study, which is due to finish in May of this year. Collectively, these outcomes demonstrate the feasibility of embedding peer-led research infrastructure within school settings.

From a participatory perspective, the outcomes of this programme align with existing literature on youth participation in research, especially those exploring the effectiveness of Young People’s Advisory Groups (YPAGs) and other co-production models.[1, 19] These approaches have consistently shown that youth involvement improves study relevance, promotes empowerment and enhances skill development.[3] However, much of the existing evidence remains concentrated on consultative or advisory roles, with limited evidence of young people being embedded across the full research lifecycle. More recent developments in co-production within adolescent mental health research have begun to address this gap. For example, structured co-analysis approaches developed by Lindsey Dewa and colleagues have demonstrated the feasibility and value of involving young people in qualitative analysis using guided frameworks, supporting shared interpretation and reflexive engagement in mental health research.[22] Such methods have since been successfully applied in broader youth mental health research contexts.[23] However, these approaches have largely remained within qualitative paradigms, where interpretative collaboration is more readily facilitated.

In contrast, the RC programme expands existing participatory models in a more comprehensive and methodologically novel direction. Specifically, it operationalises youth co-production across the entire research pipeline, including recruitment, study delivery and data collection, while also incorporating youth involvement within quantitative research processes, an area that remains underdeveloped in adolescent mental health research. Notably, this programme represents one of the few school-based participatory research models in which young people are embedded not only in study implementation but also in progressing quantitative datasets toward analysis and dissemination. This deeper level of engagement moves beyond a participatory design towards a more integrated model of co-produced quantitative research. Furthermore, the RC demonstrates that young people can actively shape empirical research processes often considered inaccessible within traditional frameworks. This includes drawing on participants’ unique insights that researchers do not have access to, such as context-specific school and peer dynamics, whilst developing transferrable skills. This positions the RC as a methodological extension of existing co-production models within adolescent mental health research.

Importantly, this expanded model of participation also underscored several practical and methodological advantages. Schools were more willing to participate when the programme provided tangible benefits for students and aligned with curriculum requirements, particularly for university applications and career progression, including for those who may otherwise lack access to such opportunities. Further, this approach enhanced school and participant involvement by placing minimal demands on teaching staff and shifting agency to the young people. Empowering students to make autonomous decisions about participation enabled RC student teams to reach their peers more quickly and efficiently than researchers alone, contributing to the collection higher data volumes within shorter timeframes. Recruitment was further accelerated through its simultaneous implementation across multiple schools, with the RC teams operating in parallel. During in-school data collection sessions, students were also more willing to take part when encouraged by familiar RC students rather than external researchers. For example, students demonstrated greater readiness to provide their data and biosamples when observing RC student teams participate themselves. Participant feedback supported this observation, with many students reporting positive experiences of the data collection sessions and expressing interest in future SCAMP involvement, including joining the YPAG and supporting further data collection activities. Although participation in more advanced stages of data analysis was variable due to time constraints, student feedback consistently highlighted gains in confidence, teamwork, research skills and insight into future academic and career pathways. This suggests that even partial involvement in quantitative research processes can yield meaningful educational and developmental benefits when appropriately designed and delivered.

Nonetheless, several challenges warrant consideration. Variations in staff continuity and the timing of initial outreach affected school engagement between years, underscoring the importance of pre-emptive, consistent communication and alignment with the school calendar. Further, although teacher-guided self-selection for the RC student teams aimed to ensure students with a genuine interest in research signed up, some students were teacher-nominated.

This introduces potential opportunity bias, whereby already high-achieving or well-known students, particularly those with whom teachers had stronger personal relationships, may have been preferentially selected for participation. Moreover, differences in how recruitment and data collection sessions were scheduled across schools, such as during lunch breaks, integrated into timetabled lessons or embedded within specific subjects, may have introduced further biases in student participation. Additionally, variation in involvement within student teams sometimes affected the pace at which deliverables were completed, as collaborative challenges were not always visible to researchers working at a distance. Future iterations should explore standardised and transparent school and student recruitment methods to promote inclusivity and ensure equitable access, place greater emphasis on clarifying student team expectations (e.g., through shared group agreements), and offer more regular check-ins and additional training. These obstacles highlight the importance of aligning research timelines with school calendars, as well as maintaining consistent communication and building trusted relationships with students and schools alike.

The success of this participatory model also has broader implications for policy and practice. Research institutions stand to benefit from adopting similar frameworks that recognise young people as co-researchers rather than passive participants. Embedding co-production models within educational-health research partnerships could enhance recruitment efficiency, improve data quality and strengthen public trust in research. Within the RC, students’ unique insights into their school environments and peer relationships substantially enriched the research process and outcomes, illustrating the value of youth-led perspectives. Crucially, the programme reinforces the principle of reciprocity as a central mechanism of engagement, where young people and schools are more likely to participate meaningfully when the research provides tangible benefits such as upskilling, and confidence and knowledge building. Whilst students received a small monetary incentive for contributing to data and biosamples as SCAMP participants, the RC team members were not remunerated for their broader research contributions. This was because non-monetary benefits (e.g., skills development, recognition and university application experience) were viewed by students as meaningful incentives for their collaboration. From a policy standpoint, the RC underscores the value of partnerships between education and health research sectors to advance youth development, health literacy and civic engagement. From a practice perspective, offering formal recognition, such as certification, academic credit as well as a minor monetary incentive for student contributions, further motivates participation, promotes equity and ensures reciprocity.

Overall, the RC demonstrates that co-developing research with young people can yield mutual benefits by advancing scientific objectives while supporting student skill development, engagement and empowerment. The programme bridged the gap between academic research and real-world settings by enabling students to shape, refine and advance research grounded in their own experiences, including generating research questions using SCAMP data and contributing to dissemination through structured feedback and training. These outcomes illustrate how the RC successfully met its objectives while delivering meaningful educational and developmental benefits for participating students. By embedding participatory research models in schools, future studies can generate richer, more relevant data whilst nurturing the next generation of researchers, informed citizens and community advocates.

## Data Availability

De-identified SCAMP data and code are available on request from.

## Supporting information

## Acknowledgements

We would like to express our gratitude to all the students, staff and schools who participated in SCAMP’s Research Challenge programme. We would like to thank all past and present SCAMP research team members for their hard work, insights and contributions. We also thank the many casual workers and interns who helped throughout the programme.

## Data availability

De-identified SCAMP data and code are available on request from.

## Author’s contributions

RBE and RT conceived the initial Research Challenge concept, whilst the programme was undertaken and further developed by NC and RT. NC, RBE, RT, LC, CS and RBS contributed to data collection sessions in schools. NC, RT, LC, CS and RBS contributed to the student research projects (including liaising with/supporting student teams, data preparation, data analysis and manuscript development). NC and RT interpreted the findings across the two years for the purpose of this manuscript. MBT, MDS, RT, LC, CS and RBS obtained further funding to support the Research Challenge beyond Year 1. NC, RT, RBS, CS, LC, RBE, MDS and MBT reviewed and edited this manuscript. All authors read and approved the final manuscript.

